# The usefulness of antigen testing in predicting contagiousness in COVID-19

**DOI:** 10.1101/2021.10.16.21265067

**Authors:** Tulio J. Lopera, Juan Carlos Alzate-Ángel, Francisco J Díaz, María T. Rugeles, Wbeimar Aguilar-Jiménez

## Abstract

Increasing the diagnostic capacity of COVID-19 (SARS-CoV-2 infection) is required to improve case detection, reduce COVID-19 expansion, and boost the world economy. Rapid antigen detection tests are cheaper and easier to implement, but their diagnostic performance has been questioned compared to RT-PCR. Here, we evaluate the performance of the Standard Q COVID-19 antigen test for diagnosing SARS-CoV-2 infection and predicting contagiousness compared to RT-PCR and viral culture, respectively. The antigen test was 100.0% specific but only 40.9% sensitive for diagnosing infection compared to RT-PCR. Interestingly, SARS-CoV-2 contagiousness is highly unlikely with a negative antigen test since it exhibited a negative predictive value of 99.9% than viral culture. Furthermore, a cycle threshold (Ct) value of 18.1 in RT-PCR was shown to be the one that best predicts contagiousness (AUC 97.6%). Thus, screening people with antigen testing is a good approach to prevent SARS-CoV-2 contagion and allow returning to daily activities.

## Introduction

Coronavirus disease 2019 (COVID-19) caused by SARS-CoV-2 has resulted in a global health crisis that requires substantial efforts worldwide to increase the diagnosis capacity and improve detection of cases by using inexpensive, easy, and rapid testing *(1)*. On the other hand, the economic re-opening requires a diagnostic test before returning to daily activities. Therefore, a negative result in a diagnostic test has become the entrance door to many countries or to other activities that involve some risk of transmission *(2)*.

Reverse transcription-polymerase chain reaction (RT-PCR) has become the standard gold test for SARS-CoV-2 detection due to its high sensitivity and specificity *(3)*. However, it has been observed that RT-PCR remains positive well beyond the period of contagiousness. Therefore, the return to daily activities is delayed, generating unnecessary restrictions for patients in the convalescent period. In these patients, viral RNA can be detected in low loads (∼Ct value > 24 in RT-PCR), and contagiousness is unlikely. For this reason, classical techniques such as viral isolation are used as a better predictor of viral infectivity. However, these methodologies are expensive, risky, and time-consuming.

In 2020 point-of-care (POC) tests, such as antigen detection, were developed and approved *(4)*. The first antigen test approved for diagnosis use in Colombia was the Standard Q COVID-19 Ag test (SD Biosensor, Republic ok Korea), which is based on immunochromatography and detects the nucleocapsid (N) antigen of SARS-CoV-2 using monoclonal antibodies *(5)*. Antigen detection tests have shown desirable diagnostic characteristics such as reasonable specificity, fast execution, and easy processing *(5,6)*.

The sensitivity and specificity of diagnostic tests are essential parameters used to guide decision-making regarding diagnostic tests. However, antigen tests are considered inferior to RT-PCR because their sensitivity for detecting SARS-CoV-2 is lower *(7,8)*, likely since the performance of antigen tests can be affected by the days of symptoms onset (DSO) and viral load during infection *(9,10)*. Likewise, the usefulness of the antigen test for predicting contagiousness remains to be determined *(11)*. Thus, we aimed to evaluate the performance of a rapid antigen detection test approved for use in Colombia using RT-PCR and viral culture as the standard gold tests for diagnosing infection and contagiousness, respectively.

## Materials and Methods

### Population

A sample size of at least 250 specimens with a ratio of positive: negative of 1:1 was calculated, anticipating a 97% specificity according to reported previously *(12,13)*, a probability of type I error of 5%, and a precision of 3%. The inclusion criteria to participate in the study were: adults (≥18 years) with suspicion of SARS-CoV-2 infection either by clinical or epidemiological criteria (clinical symptoms or recent exposure to a confirmed case, respectively). We analyzed 306 nasopharyngeal samples from 282 ambulatory subjects (some were sampled more than once as part of their follow-up) recruited at the Grupo Inmunovirología, University of Antioquia, Medellin, Colombia, between September 2020 and January 2021. We excluded pediatric patients and individuals who were unwilling to provide their written consent. The study was designed and conducted following the Declaration of Helsinki and Colombia legislation (Ministry of Health resolution 008430 of 1993). It was approved by the Ethics Committee of the Universidad de Antioquia (Act. 004, 02-04-2020). After explaining the project and clarifying doubts about the research, all included subjects signed the informed consent. The collected biological material was encoded to ensure privacy.

### Sample collection

Nasopharyngeal swabs collected on viral transport medium were obtained following the Centers for Disease Control and Prevention (CDC) recommendations *(14)*. The nasopharyngeal samples were maintained at 4°C for 4-84 hours before processing. The epidemiological and demographic data were collected from each subject filling out the official form for reporting acute respiratory infection by SARS-CoV-2 (National Institute of Health, Colombia) *(15)*.

### Antigen test

We used the Standard Q COVID-19 Ag test (SD Biosensor, Republic of Korea), following the manufacturer’s recommendations *(6)*. Briefly, 350μL of the viral transport medium were mixed for 1-10 minutes with the antigen test-lysing reagent. Then, 4 drops were added to the cassette, and after 30 minutes, the test was interpreted as positive, negative, or invalid. The antigen tests were evaluated independently by two laboratory technicians blinded to the RT-PCR and viral culture results. In case of discrepancy, the antigen test was repeated with the same sample.

### Real-time RT-PCR

Viral RNA extraction was performed from a 300 μL viral transport medium using the column-based Quick-RNA Viral Kit (Zymo Research, Orange, CA) following the manufacturer’s instructions. SARS-CoV-2 viral RNA was detected using the Luna® Universal Probe One-Step RT-qPCR Kit (New England Biolabs, MA, USA). The reaction included 7μL of viral RNA, the oligos and probe for the E gene and the conditions reported in the Berlin real-time RT-PCR protocol v2 (*16)* with a thermal modification in reverse transcription (55°C for 18 minutes) and in alignment/extension step (60°C for 30 seconds), according to the One-Step RT-qPCR Kit manufacturer’s recommendations.

In addition, human RNAse P gene transcripts were detected as internal control and evaluation of the quality of the sample, as previously recommended *(17)*. The RT-PCR reactions were carried out in a CFX-96 Biorad thermal cycler (Biorad, CA, USA). Tests were performed in parallel with a negative control (sample replaced by water) and a positive control (RNA from virus isolated at the University of Antioquia) *(18)*.

### Viral culture

Viral cultures were carried out in a biosafety level 3 laboratory (BSL-3). Approximately 100 μL of the viral transport medium was dissolved in 250 μL of DMEM culture medium supplemented with 2% fetal bovine serum and 1% Penicillin-Streptomycin. This solution was inoculated into 1 × 10^5^ Vero E6 cells monolayers in 12-well plates and incubated for 90 min at 37° C with 5% CO_2_. Subsequently, the inoculum was removed, and 1,5 mL of DMEM culture medium supplemented with 2% fetal bovine serum and 1% Penicillin-Streptomycin was added. The culture was kept at 37°C with 5% CO_2_ for 5 days. The monolayer was observed daily for a cytopathic effect (CPE; observed as rounding and detachment of infected cells) indicative of SARS-CoV-2 infection. After CPE observation, the supernatants were harvested and stored at −80°C. For confirmation purposes, some samples with or without CPE were evaluated for the presence of SARS-CoV-2 by indirect immunofluorescence *(18)* or by qRT-PCR on cell supernatants; all of these samples showed a perfect correlation between the presence of CPE and detection of the virus in the cell culture (data not shown).

### Statistical analysis

The Standard Q COVID-19 Ag test was evaluated using the sensitivity, specificity, predictive value, and likelihood ratio calculated using GrapdPad Prism (version 9, California, USA), R v4.1.0, and the Integrated Development Environment Rstudio. To explore variables associated with the result of each test, bivariate analyses were carried-out, and according to statistical (p-value <0.05) and plausibility criteria, they were included in a binary logistic regression model. Receiver Operating Characteristic (ROC) curves were constructed to discriminate the best cycle threshold (Ct) value predicting contagiousness.

## Results

### Demographic and clinical characteristics of the population

In this study, 306 nasopharyngeal samples were analyzed. All samples were blinded and codified before the analyzes. The median age of subjects was 38 years [Range 18 - 96], and 58.9% were female. Considering that the performance of diagnostic tests may fluctuate depending on DSO, we allocate the samples in four diagnostic sceneries: *i)* 108 samples came from subjects with 1-5 DSO, *ii)* 50 from individuals with 6-11 DSO, *iii)* 9 samples were from individuals with more than 11 DSO, and *iv)* 139 samples were from SARS-CoV-2-exposed subjects without symptoms (some symptomatic subjects also disclose previous close contact with a person diagnosed with COVID-19) (Figure 1).

**Figure 1:**
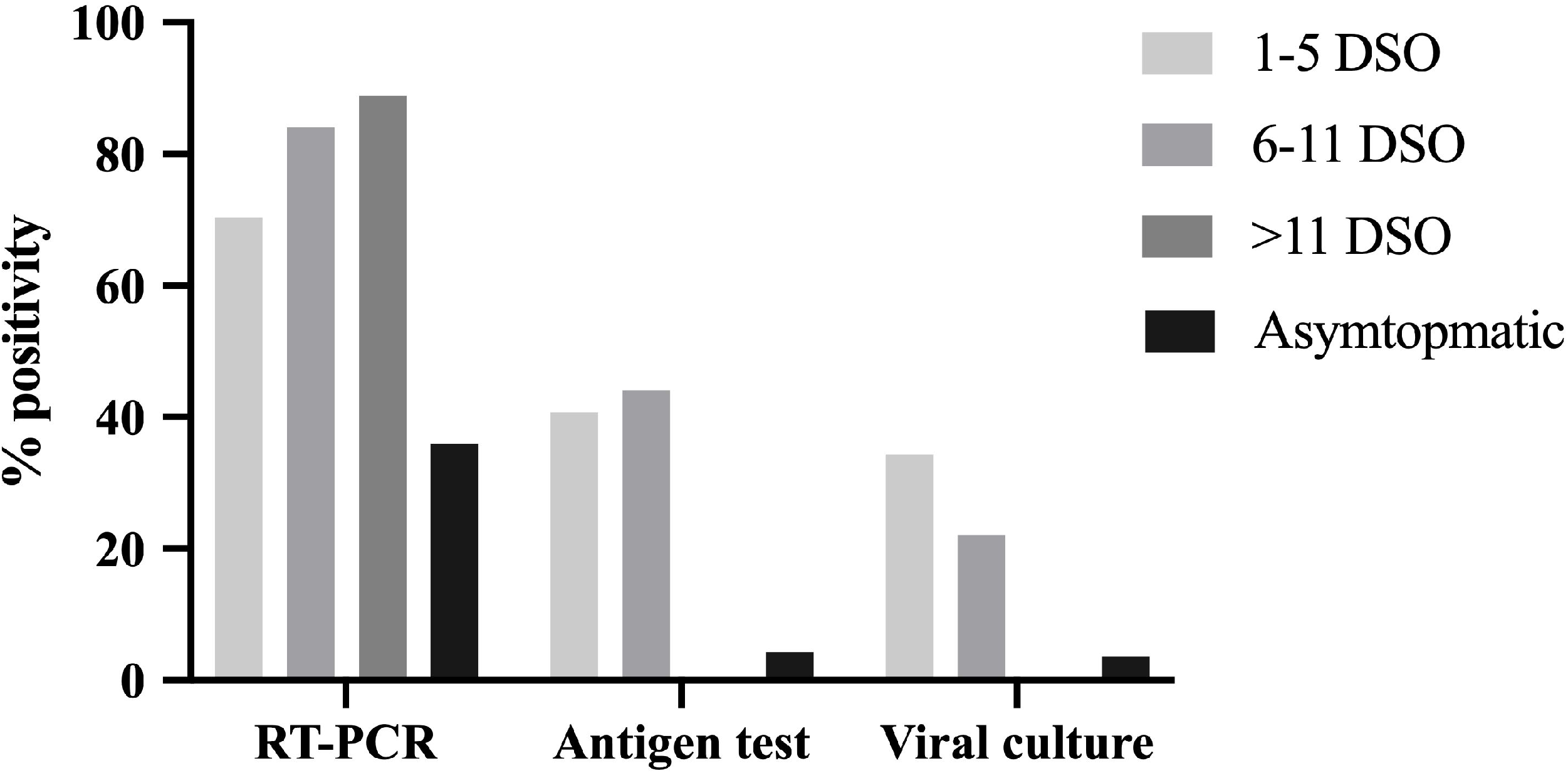
Positivity (%) of each test according to DSO or asymptomatic condition: figure shows the percentage of positive results obtained by RT-PCR, antigen test, and viral culture in patients who were on 1-5 days of symptom onset (DSO), 6-11 DSO, >11 DSO, and people who were asymptomatic individuals.

The percentage of positivity in RT-PCR was higher than 70% in individuals with symptoms but below 40% in asymptomatic ones. In contrast, less than 40% positivity by antigen test and viral culture were seen in subjects with either 1-5 or 6-11 DSO, and no positivity was observed in subjects with more than 11 DSO. As expected, the positivity in viral culture decreased as days with symptoms increased (Figure 1).

The definitive diagnosis of infection was made by RT-PCR. In total, 176 samples were SARS-CoV-2 RT-PCR-positive (57.5% positivity), most of them from outpatients who resolved COVID-19 at home, and seven were from hospitalized patients. The clinical and demographic characteristics of subjects in the study were recorded in search of associations with the result of antigen test (Table S1) and viral culture (Table S2).

### Performance of antigen test in the diagnosis of infection

People who tested positive by antigen test were older (age median [range] = 45.5 [20.0-96.0] than people tested negative (age median [range] = 37 [18.0-84.0]), and a higher proportion of them exhibited cough, fever, odynophagia, dyspnea, fatigue, conjunctivitis, headache, and anosmia or ageusia, compared to those with a negative antigen test. No differences in gender between subjects with positive or negative antigen tests were observed (Table S1).

The performance of the SARS-CoV-2 antigen test for infection diagnosis was evaluated with RT-PCR as the reference standard. From 176 RT-PCR positive specimens, 72 were positive by antigen test, revealing a low sensitivity (40.9%, confidence interval (CI) 95%= 33.6-48.6%). No negative RT-PCR sample was positive in the antigen test, demonstrating high specificity for SARS-CoV-2 infection (100% CI 95% = 96.4%-100%). Indeed, the probability of COVID-19 was high in the participants with a positive result in the antigen test (positive predictive value, PPV 100%, CI 95%= 93.7-100%). Nonetheless, the concordance between the antigen test and RT-PCR was generally low, with a kappa index of 0.37 (CI 95%= 0.29-0.44).

We observed the best performance of antigen test in samples from patients with 1-5 DSO, in which the diagnostic sensitivity was 57.9% (CI 95%= 46.0%-68.9%), the specificity was 100% (CI 95%= 86.7%-100%), and concordance with the RT-PCR using the kappa index was 0.44 (CI95%= 0.32-0.58). In patients between 6 and 11 DSO the sensitivity and specificity of the antigen test were 52.4% (CI 95%= 36.6%-67.7%) and 100% (CI 95%= 59.8%-100%), respectively. In asymptomatic subjects, the performance of the antigen test decreased, showing a poor sensitivity (12.0% CI 95%= 5.0%-25.0%), although the specificity was similar to the other categories (100% CI 95%= 94.8-100%) (Table 1).

**Table 1:** Performance of antigen test, viral culture and RT-PCR in diagnosis of infection and prediction of contagiousness of SARS-CoV-2.

| Sceneries | Sensitivity<br>(CI 95 %) | Specificity<br>(CI 95%) | VP+<br>(CI 95 %) | VP-<br>(CI 95%) | LR+<br>(CI 95 %) | LR-<br>(CI 95 %) |
| --- | --- | --- | --- | --- | --- | --- |
| **Comparison antigen test vs RT-PCR (reference)** |  |  |  |  |  |  |
| All subjects | 40.9 (33.6,48.6) | 100<br>(96.4,100) | 100<br>(93.68,100) | 55.0<br>(48.0, 61.0) | 107.31<br>(6.7, 1716.2) | 0.59<br>(0.52, 0.7) |
| 1-5 DSO | 57.9 (46.0,68.9) | 100 (86.7,100) | 1.00 (0.89,1.00) | 50.0 (38.0, 61.0) | 38.14 (2.4, 601.05) | 0.42 (0.32, 0.5) |
| 6-11 DSO | 52.4 (36.6,67.7) | 100 (59.7,100) | 100 (81.0, 100) | 28.0 (13.0, 48.0) | 9.41 (0.6, 141.4) | 0.47(0.3, 0.6) |
| Asymptomatic | 12.00 5.0,25.00) | 100 (94.8,100) | 100 (51.0, 100) | 66.0 (58.0, 74.0) | 22.94 (1.3, 398.9) | 0.88 (0.8, 0.9) |
| **Comparison antigen test vs viral culture (reference)** |  |  |  |  |  |  |
| All subjects | 96.2 (85.9,99.3) | 91.0 (87.0,94.0) | 70.0 (58.0,80.0) | 99.14 96.6,99.9) | 11.6 (7.7, 17.5) | 0.0 (0.0, 0.2) |
| 1-5 DSO | 97.0 (84.0, 99.0) | 88.0 (78.0,94.0) | 81.0 (66.0,91.0) | 98.0 (90.0, 99.0) | 8.6 (4.5, 16.6) | 0.0 (0.0, 0.2) |
| 6-11 DSO | 90.0 (57.0, 99.0) | 69.0 (52.0,82.0) | 45.0 (25.0,67.0) | 96.0 (79.0, 99.0) | 3.0 (1.8, 4.9) | 0.1 (0.0, 0.9) |
| Asymptomatic | 100 (46.3,100) | 99.0 (95.0,99.0) | 83.0 (36.0,99.0) | 100 (96.0, 100) | 134.0 (19.0,944.3) | 0.1 (0.0, 1.2) |
| **Comparison RT-PCR vs viral culture (reference)** |  |  |  |  |  |  |
| All subjects | 100 (91.0, 100) | 51.0 (45.0, 57.0) | 30.0 (23.0,37.0) | 100 (96.0, 100) | 2.1 (1.8, 2.3) | 0.0 (0.0, 0.3) |
| 1-5 DSO | 100 (88.0, 100) | 45.0 (33.0,57.0) | 48.0 (37.0,60.0) | 100 (86.0, 100) | 1.8 (1.5, 2.2) | 0.0 (0.0, 0.5) |
| 6-11 DSO | 100 (67.0, 100) | 20.0 (9.0, 36.0) | 26.0 (14.0,0.42) | 100 (59.0, 100) | 1.25 (1.1, 1.5) | 0.2 (0.0, 3.2) |
| Asymptomatic | 100 (46.0, 100) | 66.0 (57.0,74.0) | 10.0 (3.0, 22.0) | 100 (94.0, 100) | 3.0 (2.3, 3.8) | 0.1 (0.0, 1.8) |

### Positivity of Antigen test and viral culture is linked to Ct value in RT-PCR

Considering that the Ct value in RT-PCR is an indicator of viral load, we explored the distribution of Ct values between positive and negative samples by antigen test and viral culture (Figure 2). Interestingly, the Ct values among both antigen test and viral culture had a similar distribution, being lower in samples that tested positive (median Ct value [interquartile range] of 15.2 [12.1-18.1] and 13.7 [11.6-15.6] respectively) than in the negative samples (28.93 [16.7-36.6] and 27.47 [11.8-36.6] respectively, Figure 2). Indeed, out of 65 samples with a Ct<20 by RT-PCR, 62 (95.4%) were also positive by antigen test, and 51 (78.5%) were also positive by viral culture.

**Figure 2:**
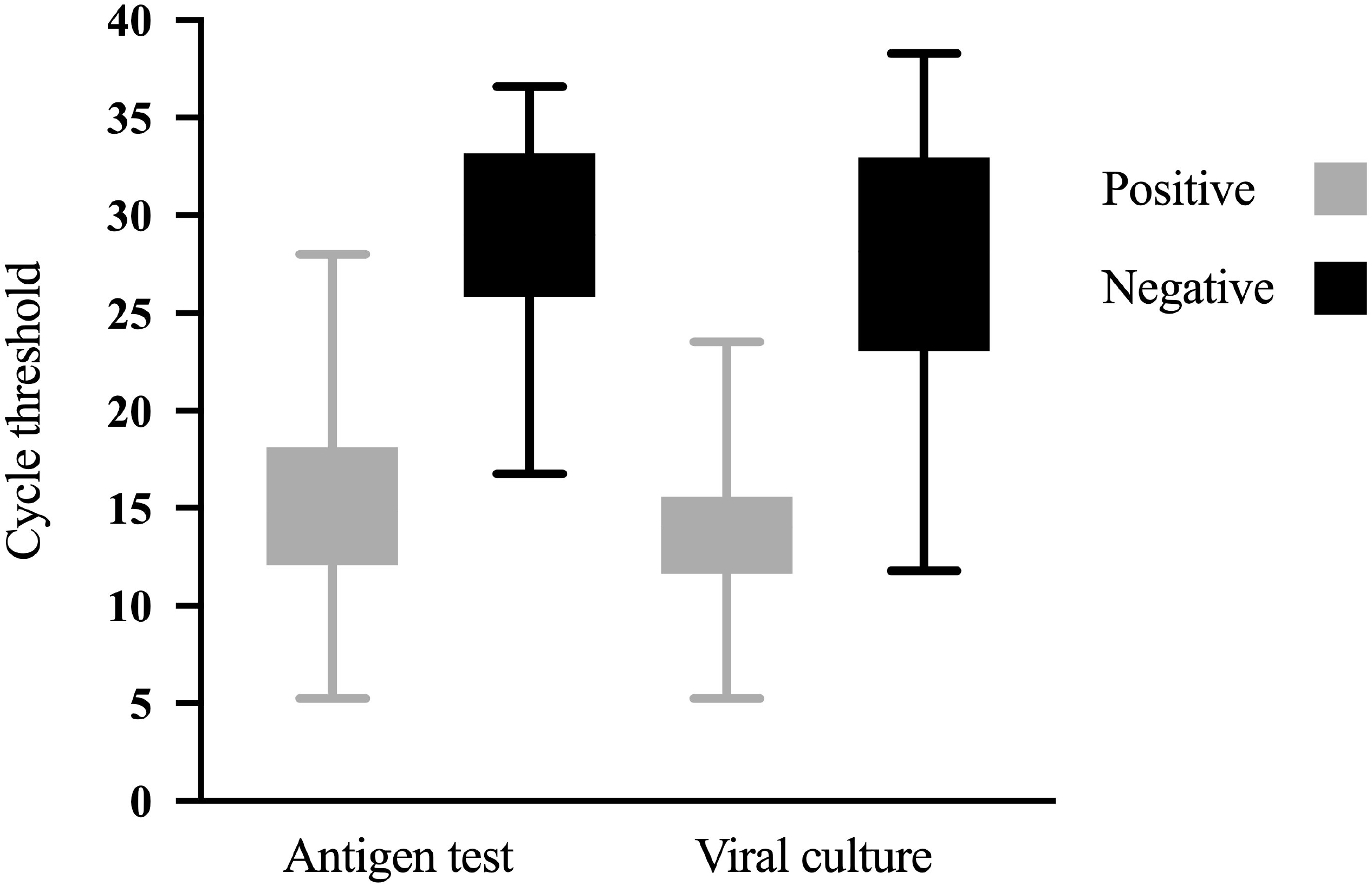
Ct values according to results in the antigen test and viral culture: Figure displays the distribution according to of cycle threshold (Ct) value obtained in RT-PCR of positive and negative results of antigen test and viral culture in patients with SARS-CoV-2 infection.

### Prediction of contagiousness

All samples were assessed by viral culture and antigen test, even when the RT-PCR was negative. SARS-CoV-2 was isolated in 53 (17,3%) of 306 samples (30% when considering only the 176 samples positive by RT-PCR). The clinical and demographic characteristics of population were recorded in search of associations with the result of viral culture, and thus its usefulness to predict contagiousness. People with a positive culture test were older (a median age [range]= 44.0 [20.0-96.0] than people without SARS-CoV-2 isolation (a median age [range]= 38.0 [18.0 – 86-0]), and had a significantly higher frequency of fever, cough, odynophagia, fatigue, dyspnea, headache and anosmia or ageusia, compared to those with a negative culture (Table S2).

We subsequently explored the usefulness of antigen tests in predicting contagiousness, as measured by viral isolation *in vitro* (Table 1). The concordance between viral culture and antigen test was acceptable, with a kappa index of 0.77 (CI95%= 0.68-0.85). Of 53 viral culture-positive samples, 51 were also positive in the antigen test, showing a high sensitivity (96.2% CI 95%= 85.9%-99.3%). In addition, we observed a high negative predictive value of 99.1% (CI 95%= 96.6%-99.9%) in predicting contagiousness (Table 1). Likewise, we found that the antigen test predicts contagiousness more accurately 1-5 DSO (kappa index of 0.82, CI95%= 0.71-0.93) than 6-11 DSO (kappa index of 0.44, CI95%= 0.21-0.67).

Asymptomatic subjects that were RT-PCR positive, usually have a low viral load (median Ct [interquartile range] = 29.6 [26.79-33.44]); therefore, most samples were negative by antigen test (95.6%) and viral culture (96.4%). Nevertheless, in this group, the sensitivity of the antigen test for contagiousness was high (100% [CI 95%= 46.29%-100%]) (Table 1).

We carried out a multiple logistic regression model (Table S3) with variables such as the Ct value in RT-PCR, and those showing a crude association with antigen test or viral culture results (Tables S1 and S2). Although positivity in antigen test was associated with positivity in viral culture [crude OR=134.79 (30.45,596.64)], the association was not statistically significant after covariates adjustment [adjusted OR=5.79 (0.5,66.85)] (Table S3). In contrast, the categorized Ct value was associated with the positivity in viral culture after adjustment. Indeed, the samples with a Ct<20 and <15 had adjusted ORs of 25.07 (2.27, 277.23) and 290.45 (17.19, 4907.16)] respectively (Table S3).

### Predicting contagiousness by RT-PCR

According to the previous findings, the RT-PCR Ct value, as an indicator of viral load, could help predict infection. Therefore, we analyze the Ct value to find the value that best predicts a positive result in viral culture. An analysis of a receiver operating characteristic (ROC curve) was performed, obtaining a Ct value of 18.1 as the best predictor of viral culture result, with an area under the curve (AUC) of 97.6 % (95% CI = 95.6% - 99.5%) (Figure 3). Our results also showed that the highest Ct value where a viral isolate was obtained was 23.5 (Figure 2).

**Figure 3.**
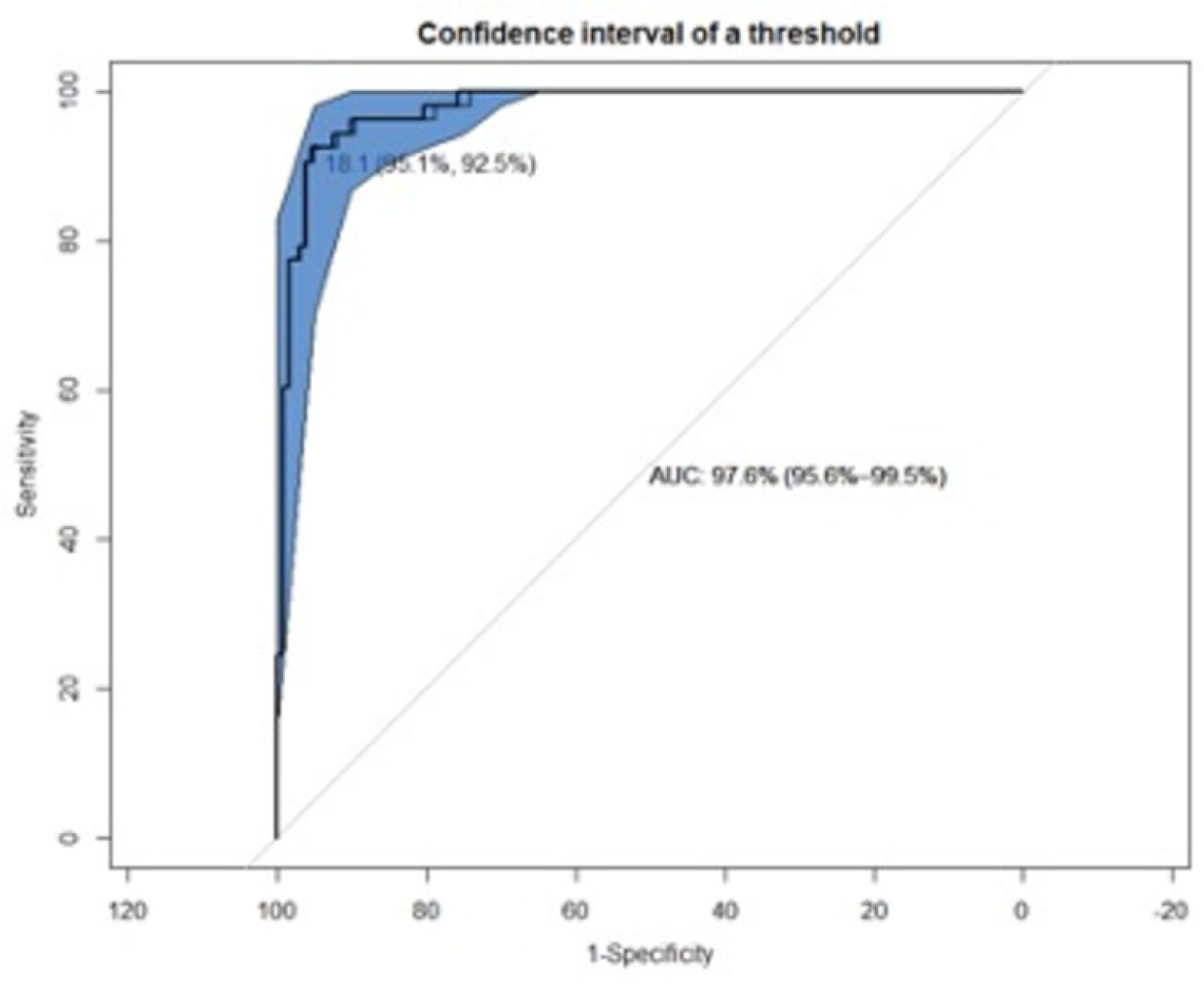
ROC Curve and AUC to calculate the discriminatory Ct value in contagiousness. The Receiver operating characteristic (ROC) curve shows the Ct value where sensitivity and specificity are higher than 95% to predict the contagiousness using viral culture as reference. Area Under the Curve (AUC) 97.6% (95.6%-99.5%).

## Discussion

We evaluated the Standard Q antigen test performance to diagnose SARS-CoV-2 infection and predict contagiousness. This rapid and simple antigen test is widely used in Colombia and other countries, especially in high prevalence populations; its sensitivity has varied widely across different evaluations *(13,16,19–22)*. In this study, the sensitivity for infection diagnosis in all samples was low, 40.9% (CI 95%= 33.6,48.6%), much lower than the 84% sensitivity reported by the manufacturer *(6)*. Nevertheless, other authors have reported significant variation in sensitivity ranging between 42.9% *(23)*, 70.7% *(24)*, and 98.3% *(16)*.

This variability seems to be only partially explained by the DSO at sampling because the sensitivity of antigen test in subjects with 1-5 DSO increases up to 57%, whereas in subjects with 6-11 DSO the sensitivity was 52%. Indeed, other studies have reported similar sensitivity values in subjects with ≤14 DSO (55.4%) *(22)*. On the other hand, our sampling population consists mostly of outpatients, which usually have mild disease and consequently lower viral load. Actually, we observed similar sensitivity compared to 63% reported in patients with the same disease spectrum *(25)*. Indeed, when we evaluated the performance of the antigen test compared to the Ct in RT-PCR, the proportion of positive antigen test was 62/65 (95.4%) in samples with Ct<20, in concordance with other reports indicating around 95% sensitivity of antigen test in samples with high viral load (Ct less than 22.5) *(26,27)*.

Although the sensitivity of the antigen test was low, its high specificity and high positive predictive value indicate that a positive result in this test provides reliable evidence of SARS-CoV-2 infection; conversely, a negative result does not rule out infection. However, additional testing in subjects with a negative antigen test could be unnecessary if we are interested in detecting patients who are shedding viruses and therefore are a threat to those close to them.

Remarkably, the antigen test and viral culture performance were similar; in both cases, the distribution of Ct values had a median <20 in positive results. Whereas there were no specific symptoms typical in COVID-19 or DSO related to the probability of viral isolation, the antigen test showed a specificity of 91% to predicts viral infectivity; from the 306 samples, only 2 were discordant between antigen test and viral culture results. In addition, our findings are consistent with studies such as Pekosz *et al (28)*, who reported that the antigen tests correlate with viral culture of SARS-CoV-2 in 27/28 samples assessed. Yamayoshi *et al (26)*, in samples with a Ct value <22.5 found that antigen test result was similar to viral isolation in culture (11/11 were positive in antigen test and 8/11 in viral culture). Therefore, these results suggest that an antigen test is a promising tool to discharge patients from quarantine because of the low probability of transmitting the virus.

Although viral culture is the most obvious method to assess contagiousness, it is complex, expensive, and requires particular biosafety conditions. The Ct value of RT-PCR, despite the usefulness in the prediction of contagiousness in samples with high viral load, is intrinsically variable *(25,29,30)*. The variation in Ct can be due to the conditions of the RT-PCR reaction, the fluorescence threshold, the fluorochrome used, the gene target, and the virus lineage *(10)*. Antigen test, which targets the more conserved nucleocapsid protein, is potentially a more consistent and cost/benefit method of assessing contagiousness.

Although some authors have reported viral isolation in culture up to Ct value close to 32 *(25,31,32)*, most studies have associated viral isolation with Ct 18-24 (5.4 – 7.0 log RNA copies) *(33,34)*. Our study indicates that the Ct value that best predicts the result in viral culture is 18.1, with a progressive decrease in the possibility of contagiousness above this Ct value. Hence, we propose that a Ct value higher than 23.5 could be a safe threshold of contagiousness since it was the highest Ct in which we could isolate the virus in culture.

In this study, the time of exposure was not controlled since it was reported by each individual, considered from the last contact with someone with a COVID-19 confirmed diagnosis. Due to this, many individuals did not know precisely the time of exposure to the virus. Additionally, the data correspond to a study of diagnostic tests in which the prevalence of infection is given by the proportion of infected and uninfected subjects who participated in the study, which could not be an exact reflection of the actual prevalence in the population. Further studies should include a serial sampling of the participants in other prevalence scenarios to improve the scope of the results.

## Conclusion

This study demonstrated that the Standard Q COVID-19 antigen test has excellent agreement with viral culture, indicating that it can be used as a marker of contagiousness. Due to its high positive predictive value in situations of a high prevalence of infection, positive results do not require confirmation with another test. Likewise, its high negative predictive value for contagiousness makes it possible to use this test as a criterion to discharge patients in isolation and screen people moving into environments that could facilitate the transmission of the virus. Additionally, we found that 18.1 is the Ct value in RT-PCR that best predicts the possibility of *in vitro* viral isolation, related to a higher probability of contagion and viral transmission.

## Data Availability

All data produced in the present study are available upon reasonable request to the authors

## Acknowledgments

The authors thank the patients and volunteers who kindly participated in this study. We also thank the clinicians who support the recruitment of patients and laboratory assistants who contributed to the sampling.

## Author Contributions

“Conceptualization, T.J.L, F.J.D, J.C.A, M.T.R, and W.A.J.; methodology, T.J.L, and W.A.J.; formal analysis, J.C.A, T.J.L, and W.A.J; writing—original draft preparation, T.J.L, J.C.A, and W.A.J; writing—review and editing, M.T.R, J.C.A., F.J.D., and W.A.J. All authors have read and agreed to the published version of the manuscript.”

## References

1. Cheng MP, Papenburg J, Desjardins M, Kanjilal S, Quach C, Libman M, et al. Diagnostic Testing for Severe Acute Respiratory Syndrome-Related Coronavirus 2: A Narrative Review. Ann Intern Med. 2020 Jun;172(11):726–34.

2. Oztig LI, Askin OE. Human mobility and coronavirus disease 2019 (COVID-19): a negative binomial regression analysis. Public Health. 2020 Aug; 185:364–7.

3. Organization WH. Diagnostic testing for SARS-CoV-2: interim guidance, 11 September 2020. World Health Organization; 2020.

4. Organization WH. Antigen-detection in the diagnosis of SARS-CoV-2 infection using rapid immunoassays: interim guidance, 11 September 2020 [Internet]. Geneva PP -Geneva: World Health Organization; Available from: https://apps.who.int/iris/handle/10665/334253

5. Mercado Reyes M et al. Validación secundaria y verificación del desempeño de la prueba. 2020;(293):1–7.

6. SD, BIOSENSOR. STANDARDTM Q COVID-19 Ag Test. 2020.

7. Corman VM, Haage VC, Bleicker T, Schmidt ML, Mühlemann B, Zuchowski M, et al. Comparison of seven commercial SARS-CoV-2 rapid point-of-care antigen tests: a single-center laboratory evaluation study. The Lancet Microbe. 2021;5247(21):1–9.

8. Dinnes J, Deeks JJ, Adriano A, Berhane S, Davenport C, Dittrich S, et al. Rapid, point-of-care antigen and molecular-based tests for diagnosis of SARS-CoV-2 infection [Internet]. Vol. 8, The Cochrane database of systematic reviews. Test Evaluation Research Group, Institute of Applied Health Research, University of Birmingham, Birmingham, UK.; 2020. p. CD013705. Available from: http://europepmc.org/abstract/MED/32845525

9. Linares M, Pérez-Tanoira R, Carrero A, Romanyk J, Pérez-García F, Gómez-Herruz P, et al. Panbio antigen rapid test is reliable to diagnose SARS-CoV-2 infection in the first 7 days after the onset of symptoms. J Clin Virol Off Publ Pan Am Soc Clin Virol. 2020 Dec; 133:104659.

10. Rabaan AA, Tirupathi R, Sule AA, Aldali J, Mutair AA, Alhumaid S, et al. Viral Dynamics and Real-Time RT-PCR Ct Values Correlation with Disease Severity in COVID-19. Vol. 11, Diagnostics. 2021.

11. Meyerowitz EA, Richterman A, Gandhi RT, Sax PE. Transmission of SARS-CoV-2: A Review of Viral, Host, and Environmental Factors. Ann Intern Med. 2021 Jan;174(1):69– 79.

12. ICMR. Advisory on Use of Rapid Antigen Detection Test for COVID-19. 2020;1– Available from: https://www.icmr.gov.in/pdf/covid/strategy/Advisory_for_rapid_antigen_test14062020.pdf

13. Chaimayo C, Kaewnaphan B, Tanlieng N, Athipanyasilp N, Sirijatuphat R, Chayakulkeeree M, et al. Rapid SARS-CoV-2 antigen detection assay in comparison with real-time RT-PCR assay for laboratory diagnosis of COVID-19 in Thailand. Virol J [Internet]. 2020;17(1):1–7. Available from: https://doi.org/10.1186/s12985-020-01452-5

14. CDC. Interim Guidelines for Collecting and Handling of Clinical Specimens for COVID-19 Testing: Updated Feb. 26, 2021. Centers Dis Control Prev [Internet]. 2021;1–6. Available from: https://www.cdc.gov/coronavirus/2019-ncov/lab/guidelines-clinical-specimens.html

15. Instituto Nacional de Salud. Ficha de notificación individual – Infección respiratoria aguda por virus nuevo. 2020;2019.

16. Corman VM, Landt O, Kaiser M, Molenkamp R, Meijer A, Chu DKW, et al. Detection of 2019 novel coronavirus (2019-nCoV) by real-time RT-PCR. Eurosurveillance [Internet]. 2020;25(3). Available from: https://www.eurosurveillance.org/content/10.2807/1560-7917.ES.2020.25.3.2000045

17. Palacio Rua K, García Correa JF, Aguilar-Jiménez W, Afanador Ayala C, Rugeles MT, Zuluaga AF. Validación de una técnica de PCR dúplex usando el gen E y RNasa P para el diagnóstico de SARS-CoV-2. Enferm Infecc Microbiol Clin [Internet]. 2021; Available from: https://www.sciencedirect.com/science/article/pii/S0213005X21000197

18. Díaz FJ, Aguilar-Jiménez W, Flórez-Álvarez L, Valencia G, Laiton-Donato K, Franco-Muñoz C, et al. Aislamiento y caracterización de una cepa temprana de SARS-CoV-2 durante la epidemia de 2020 en Medellín, Colombia. Biomédica [Internet]. 2020;40(Supl. 2):148–58. Available from: https://revistabiomedica.org/index.php/biomedica/article/view/5834

19. Homza M, Zelena H, Janosek J, Tomaskova H, Jezo E, Kloudova A, et al. Five antigen tests for sars-cov-2: Virus viability matters. Viruses. 2021;13(4):1–9.

20. Jegerlehner S, Suter-Riniker F, Jent P, Bittel P, Nagler M. Diagnostic accuracy of a SARS-CoV-2 rapid antigen test in real-life clinical settings. Int J Infect Dis IJID Off Publ Int Soc Infect Dis. 2021 Aug; 109:118–22.

21. Hayer J, Kasapic D, Zemmrich C. Real-world clinical performance of commercial SARS-CoV-2 rapid antigen tests in suspected COVID-19: A systematic meta-analysis of available data as of November 20, 2020. Int J Infect Dis IJID Off Publ Int Soc Infect Dis. 2021 Jul; 108:592–602.

22. Hirotsu Y, Maejima M, Shibusawa M, Nagakubo Y, Hosaka K, Amemiya K, et al. Comparison of automated SARS-CoV-2 antigen test for COVID-19 infection with quantitative RT-PCR using 313 nasopharyngeal swabs, including from seven serially followed patients. Int J Infect Dis IJID Off Publ Int Soc Infect Dis. 2020 Oct; 99:397–402.

23. Korenkov M, Poopalasingam N, Madler M, Vanshylla K, Eggeling R, Wirtz M, et al. Evaluation of a Rapid Antigen Test To Detect SARS-CoV-2 Infection and Identify Potentially Infectious Individuals. J Clin Microbiol. 2021;59(9).

24. Krüger LJ, Gaeddert M, Köppel L, Brümmer LE, Gottschalk C, Miranda IB, et al. Evaluation of the accuracy, ease of use and limit of detection of novel, rapid, antigen-detecting point-of-care diagnostics for <em= SARS-CoV-2</em= medRxiv [Internet]. 2020 Jan 1;2020.10.01.20203836. Available from: http://medrxiv.org/content/early/2020/10/04/2020.10.01.20203836.abstract

25. La Scola B, Le Bideau M, Andreani J, Hoang VT, Grimaldier C, Colson P, et al. Viral RNA load as determined by cell culture as a management tool for discharge of SARS-CoV-2 patients from infectious disease wards. Eur J Clin Microbiol Infect Dis. 2020;39(6):1059–61.

26. Yamayoshi S, Sakai-Tagawa Y, Koga M, Akasaka O, Nakachi I, Koh H, et al. Comparison of rapid antigen tests for covid-19. Viruses. 2020;12(12):1–8.

27. Hirotsu Y, Maejima M, Shibusawa M, Nagakubo Y, Hosaka K, Amemiya K, et al. Comparison of automated SARS-CoV-2 antigen test for COVID-19 infection with quantitative RT-PCR using 313 nasopharyngeal swabs, including from seven serially followed patients. Int J Infect Dis [Internet]. 2020; 99:397–402. Available from: https://doi.org/10.1016/j.ijid.2020.08.029

28. Pekosz A, Parvu V, Li M, Andrews JC, Manabe YC, Kodsi S, et al. Antigen-Based Testing but Not Real-Time Polymerase Chain Reaction Correlates with Severe Acute Respiratory Syndrome Coronavirus 2 Viral Culture. Clin Infect Dis. 2021;21205(Xx):1–6.

29. Singanayagam A, Patel M, Charlett A, Lopez Bernal J, Saliba V, Ellis J, et al. Duration of infectiousness and correlation with RT-PCR cycle threshold values in cases of COVID-19, England, January to May 2020. Euro Surveill Bull Eur sur les Mal Transm = Eur Commun Dis Bull. 2020 Aug;25(32).

30. Bhat TA, Kalathil SG, Bogner PN, Blount BC, Goniewicz ML, Thanavala YM. An Animal Model of Inhaled Vitamin E Acetate and EVALI-like Lung Injury. N Engl J Med. 2020;382(12):1175–7.

31. Wölfel R, Corman VM, Guggemos W, Seilmaier M, Zange S, Müller MA, et al. Virological assessment of hospitalized cases of coronavirus disease 2019. medRxiv. 2020;

32. Jeong HW, Kim S, Kim H, Kim Y, Kim JH, Cho JY, et al. Viable SARS-CoV-2 in various specimens from COVID-19 patients. 2020;(January).

33. L’Huillier AG, Torriani G, Pigny F, Kaiser L, Eckerle I. Culture-Competent SARS-CoV-2 in Nasopharynx of Symptomatic Neonates, Children, and Adolescents. Emerg Infect Dis. 2020;26(10):2494–7.

34. Yu F, Yan L, Wang N, Yang S, Wang L, Tang Y, et al. Quantitative Detection and Viral Load Analysis of SARS-CoV-2 in Infected Patients. Clin Infect Dis [Internet]. 2020 Jul 28;71(15):793–8. Available from: https://pubmed.ncbi.nlm.nih.gov/32221523

